# Anterior middle cingulate cortex gamma-aminobutyric acid level is elevated in children with both familial and prenatal alcohol exposure-associated attention deficit hyperactivity disorder

**DOI:** 10.64898/2026.05.25.26354065

**Authors:** Jeffry R. Alger, Ishika Gupta, Lea Farkouh, Jennifer Korthas, Ankita Shah, Arjun Silverberg, Noriko Salamon, Benjamin N. Schneider, Shantanu H. Joshi, Mary J. O’Connor, Joseph O’Neill

## Abstract

**Background:** Prior neuroimaging suggests brain differences between children with attention deficit hyperactivity disorder due to prenatal alcohol exposure (ADHD+PAE) and non-exposed children with ADHD due to other, e.g., familial, causes (ADHD-PAE). There has been interest in regional brain levels of gamma-aminobutyric acid (GABA) and glutamate (Glu) measured *in vivo* with magnetic resonance spectroscopy (MRS) as possible indicators of local inhibitory, respectively, excitatory activity in ADHD. For the first time, we report here a comparison of GABA and Glu in ADHD+PAE vs. ADHD-PAE.

**Methods:** At 3 T, we used J-difference-edited single-voxel MRS to assay GABA and Glu in 28 children with ADHD+PAE, 20 with ADHD-PAE, and 28 typically developing (TD) controls, all aged 8-14 years. MRS was sampled from midline anterior middle cingulate cortex (aMCC), the “cognitive cingulate” considered functionally relevant to ADHD. Spectra were fit with custom software, including a unique technique for isolating the GABA signal from the confounding macromolecular baseline (MMBL).

**Results:** aMCC GABA was higher in ADHD+PAE and ADHD-PAE than in TD. GABA increased with age in TD, but not in ADHD+PAE or ADHD-PAE. Similar effects were observed for the ratios GABA/Glu and GABA/Glx. For GABA+MMBL (GABA+) these effects were not seen, rather GABA+ and MMBL increased with age for the ADHD+PAE group only. No significant effects were found for Glu or Glx.

**Conclusions:** GABA in the aMCC does not distinguish the two etiologies of ADHD, rather elevated GABA that follows an abnormal developmental appears to be common to both. High GABA may reflect increased inhibition of the aMCC impairing its cognitive functions. GABA+ results in ADHD may not tract reliably with underlying GABA values. Negative results for Glu and Glx should be reexamined at shorter echo-times.

## INTRODUCTION

Prenatal alcohol exposure (PAE) is more prevalent than many clinicians and researchers realize (Lange et al, 2017; May et al., 2018). It is also an important, often unrecognized, etiology of attention deficit hyperactivity disorder (ADHD; Fryer et al., 2007, O’Connor, 2014, San Martin Porter et al., 2019), the most common neurodevelopmental disorder in childhood (Yang et al., 2022). In practice, children with ADHD associated with PAE (“ADHD+PAE”) are often misdiagnosed as unexposed children whose ADHD is due to other, often familial, causes (“ADHD-PAE”; Mattson et al. 2013; O’Connor, 2014; Young et al., 2016). Misdiagnosis can have consequences. Conventional psychostimulant therapy may be less effective (Doig et al., 2008; Infante et al., 2011; Oesterheld et al., 1998; Snyder et al., 1997; Young et al., 2016) and induce side effects at higher rates (Coe et al., 2001; Ji & Findling, 2016; Ipsiroglu et al., 2015) in ADHD+PAE than ADHD-PAE. Additionally, for many children with ADHD-PAE, symptoms resolve as they age into adolescence and adulthood (Jensen et al., 2007), while people with ADHD+PAE more frequently develop “secondary disabilities”, long-term adverse outcomes (O’Connor & Paley, 2009; Streissguth et al., 1997), including school and legal trouble, substance abuse, mental illness, and suicide (Burd et al., 2007; Fast & Conry, 2004; O’Connor, 2014; O’Connor & Paley, 2009; O’Connor et al., 2019), linked to persistent problems in attention and executive function, behavioral and emotional regulation, and adaptive function (O’Connor, 2014; Streissguth et al., 1997; Young et al., 2016).

Conversely, occult PAE is well represented in clinical ADHD populations (Coles, 2001), but few studies of ostensible ADHD-PAE screen for PAE. Thus, unintended mixing of PAE and non-PAE subjects in studies of ADHD may add untoward variability and cloud interpretation of results. Thus, distinguishing ADHD+PAE is important for both research and clinical practice.

But the differential diagnosis of ADHD+PAE from ADHD-PAE can be challenging. The key PAE criterion of maternal alcohol consumption during pregnancy often cannot be established as many children with PAE are adopted or in foster care with the birth mother unavailable (Chernoff et al., 1994). Further, many birth mothers are understandably reluctant to admit alcohol consumption while pregnant (CDC, 2004). Interest, therefore, remains in developing objective diagnosis with the help of neuroimaging. In our recent work (Alger et al. 2021; Gupta et al., submitted; Kilpatrick et al. 2021,2022; O’Neill et al. 2019,2022), we have applied structural MRI, diffusion tensor imaging (DTI), resting-state fMRI (rsfMRI), and proton magnetic resonance spectroscopy (MRS) in pursuit of this question and of better understanding of the neurobiological bases of ADHD in both PAE and non-PAE etiologies. We have discovered a number of brain imaging endpoints that differ between ADHD+PAE and ADHD-PAE in children and even allow single-subject classification. This report concerns a further such effort using MRS.

In the last 30 years, a method known formally as J-difference-edited single-voxel MRS, informally, Mescher-Garwood point-resolved spectroscopy—“MEGA-PRESS” (Mescher et al., 1998; Peek et al., 2023), has been widely used to measure the (approximate) level of gamma-aminobutyric acid (GABA) non-invasively in the living human brain using MRS. J-difference-editing uses radiofrequency pulses to selectively perturb the hydrogen nuclei within the GABA molecule in a way that renders the GABA nuclear magnetic resonance near 3.0 parts per million (ppm) visible in relative isolation; in conventional MR spectra, the 3.0-ppm peak is strongly overlapped by stronger neighboring signals produced by other brain metabolites. The method does not use ionizing radiation and is therefore safe for studies of the developing human brain. Use of J-difference-editing for semi-quantitative assessment of brain GABA level has been widely used in a number of contexts (Schür et al., 2016), including investigation of ADHD (Bollmann et al., 2015; Edden et al., 2012; Ende et al. 2018; Hai et al., 2020; Harris et al., 2021; Kahl et al., 2022; Pang et al., 2023; Solleveld et al., 2017). J-difference-editing used to assay GABA simultaneously measures the level of glutamate (Glu) in the same volume-of-interest. Given that GABA is a principal inhibitory neurotransmitter and Glu is a principle excitatory neurotransmitter in the brain, combined assessment of GABA and Glu offers the opportunity to evaluate the hypothesis that an inhibitory-excitatory imbalance may underlie ADHD+PAE and/or ADHD-PAE, and perhaps provide a means to distinguish the two. It has been suggested that abnormal cortical GABA is central to ADHD and that GABAergic mechanisms may be useful in development of future drugs for ADHD (Ferranti et al. 2024). The present study, is, to our knowledge, the first use of J-difference-editing to compare brain GABA and Glu between well-characterized cohorts of children with ADHD+PAE and ADHD-PAE. In addition to the two ADHD groups, typically developing (TD) children were evaluated.

We chose to measure GABA using J-difference-editing in the anterior middle cingulate cortex (aMCC), also called “dorsal anterior cingulate cortex—dACC”. A major general function of the aMCC is described as comparison of aversion (pain, discomfort, ennui) and reward in approach-avoidance behavior in the context of motor response selection (Vogt 2009,2016). One reason this is relevant to ADHD is that people with ADHD often have difficulty concentrating on boring or unpleasant (aversive) tasks and are easily distracted towards emotionally more appealing (rewarding) activities. Based on convergent neuroimaging, preclinical and other findings, dysfunction of the aMCC, referred to as the “cognitive cingulate” (Bush et al., 2000), has long been implicated in ADHD (Bush, 2011; Bush et al., 2005; Vogt, 2016,2019). The aMCC is also rich in dopaminergic afferents and receptors (Vogt 2016) and dopaminergic stimulants are first-line treatments for ADHD. Given that its deep cerebral location also affords clean spectra, the aMCC is a good site to investigate ADHD GABA.

One limitation of the GABA J-difference-edited single-voxel MRS method, as usually practiced, is that it does not perfectly isolate the GABA 3.0 ppm signal from nearby overlapping signals (Mescher et al., 1998) produced by arginine (Arg) and lysine (Lys) amino acid residues that are constituents of brain macromolecules. Arg and Lys have nuclear quantum mechanical properties that closely mimic those of GABA. This confound has encouraged use of the notation “GABA+” to report measurement of the combined levels of GABA and its confounding macromolecular baseline (MMBL). The present work statistically models the macromolecular contribution to the GABA+ signal in order to quantitate GABA and MMBL separately (Alger et al., 2019). This is a unique aspect of the present contribution.

## MATERIALS AND METHODS

### Participants

Participants were recruited and assessed as in our prior work (Alger et al. 2021; Kilpatrick et al. 2021, 2022; O’Neill et al. 2021). Briefly, recruitment sources included FASD organizations, national websites, other pediatric studies, and physician referrals. Candidate participants were screened by telephone. Eligible participants underwent demographic and medical history, diagnostic and clinical interviews, and mock scanning, then actual MRI scanning. Children taking stimulants were asked to be off medication for at least 24 hr prior to testing and scanning. The UCLA Institutional Review Board approved all procedures. All parents gave informed consent; all children gave assent and were compensated.

Seventy-six children between 8 and 14 years of age with full-scale IQ (FSIQ) ≥70 (WASI-II; Wechsler, 2011) participated in the study (Table 1). The participants were divided into three groups: children with attention deficit hyperactivity disorder and confirmed prenatal alcohol exposure (ADHD+PAE; n = 28), children with ADHD and no evidence of prenatal alcohol exposure (ADHD–PAE; n = 20), and typically developing controls (TD; n = 28). These were the final numbers analyzed after exclusions and quality assurance of MRS data (see below). Children in the ADHD+PAE group had to meet criteria for fetal alcohol syndrome (FAS), partial fetal alcohol syndrome (pFAS), or alcohol-related neurodevelopmental disorder (ARND) using the modified Institute of Medicine (IOM) criteria (Hoyme et al., 2016; see O’Connor et al., 2019). Prenatal exposure to alcohol and other teratogens was assessed using the Health Interview for Women (HIW; O’Connor & Kasari, 2000) or the Health Interview for Adoptive and Foster Parents (HIAFP; Quattlebaum & O’Connor, 2013).

**Table 1.** Demographic and clinical variables of final sample.

|  | ADHD+PAE | ADHD-PAE | TD |
| --- | --- | --- | --- |
| **N** | 28 | 20 | 28 |
| **Sex**, male/female | 16/12 | 14/6 | 13/15 |
| **Age**, years | 9.6±1.4<sup>*,†††</sup> | 10.5±1.4 | 11.0 ±1.6 |
| **Full-scale IQ** | 98.2±13.6<sup>*,†††</sup> | 108.5±12.8<sup>†</sup> | 115.3 ±17.2 |
| **Conners Inattention** | 81.8±9.3<sup>***,†††</sup> | 82.4±9.4<sup>†††</sup> | 48.2±10.7 |
| **Conners Hyperactivity** | 83.0±9.5<sup>***,†††</sup> | 79.9±13.3<sup>†††</sup> | 49.1±10.5 |
| **Concurrent Medications:** |  |  |  |
| None | 12 | 17 | 28 |
| Methylphenidate | 2 | 3 | 0 |
| Amphetamines | 6 | 0 | 0 |
| Atomoxetine | 2 | 0 | 0 |
| Guanfacine | 5 | 0 | 0 |
| Clonidine | 1 | 0 | 0 |
| Fluoxetine | 3 | 1 | 0 |
| Risperidone | 2 | 1 | 0 |
| Aripiprazole | 2 | 0 | 0 |
| Lamotrigine | 1 | 0 | 0 |
| Valproate | 1 | 0 | 0 |
| Oxcarbazepine | 1 | 0 | 0 |
| **Prenatal Exposures:** |  |  |  |
| None | 0 | 20 | 28 |
| Ethanol | 28 | 0 | 0 |
| Tobacco | 2 | 0 | 0 |
| Cannabis | 5 | 0 | 0 |
| Stimulants | 9 | 0 | 0 |
| Opioids | 2 | 0 | 0 |
| Barbiturates | 1 | 0 | 0 |
| Cocaine | 1 | 0 | 0 |
| Unspecified | 1 | 0 | 0 |

ADHD+PAE=attention deficit hyperactivity disorder likely due to prenatal alcohol exposure; ADHD-PAE=attention deficit hyperactivity disorder without prenatal alcohol exposure, likely due to genetics; TD=typically developing control. Conner Inattention—Parent Conner Inattention T-Score; Conner Hyperactivity—Parent Conner Hyperactivity/Impulsivity T-Score; \* $p < 0.05$ , \*\*\* $p < 0.001$ vs. ADHD-PAE; <sup>†</sup> $p < 0.05$ , <sup>†††</sup> $p < 0.001$ vs. TD.

Criteria for alcohol exposure included >6 drinks/week for ≥2 week and/or ≥3 drinks on ≥2 occasions including prior to and following pregnancy recognition. For adoptive/fostered participants, data regarding prenatal exposure to alcohol and other teratogens were obtained via birth, medical, or adoption records, or by reliable informants. This approach is considered acceptable for establishing PAE by the scientific community (CDC, 2004). Children in the ADHD groups met DSM-5 criteria for ADHD, any subtype, according to the Schedule for Affective Disorders and Schizophrenia for School-Aged Children Parent Version (K-SADS P; Kaufman et al., 1997, Townsend et al., 2020). Children in the ADHD-PAE group had to have one or more first-degree relatives with diagnosed ADHD to support a probable genetic etiology of ADHD and exposure to < 2 standard drinks (1.20 oz absolute alcohol) throughout gestation. Children in the TD group were required to have no first-degree relative with diagnosed ADHD and exposure to <2 standard drinks throughout gestation. Severity of ADHD was assessed by Parent ratings on the Conners 3 Behavior Rating Scale (Conners, 2008).

The three diagnosis groups (ADHD+PAE, ADHD-PAE and TD) were imperfectly matched for gender and age (Table 1). No between-group difference in gender reached significance ( χ^2^-Test, all *p* > 0.05), but the ADHD+PAE sample was significantly younger than both the ADHD-PAE (Kruskal-Wallis Test *H* = 5.9, *p* = 0.015) and TD (*H* = 11.9, *p* = 0.001) samples. Both age and gender were included in the model when examining effects on metabolite levels (see below). The ADHD+PAE group also exhibited lower FSIQ than the ADHD–PAE (*p* < 0.011) and TD (*p* < 0.001) groups. Nonetheless, FSIQ was not included as a covariate since reduced IQ is a core feature of fetal alcohol spectrum disorders, and covarying for IQ could remove variance that is intrinsic to the condition. Medication use differed across the groups, with greater medication use in the ADHD+PAE group, lower use in the ADHD–PAE group, and no medication use in the TD group. Medication use was not included as a covariate, as it reflects the clinical phenotype of the population.

### MRI and MRS acquisition

Prior to MRI, participants underwent extensive desensitization to the MRI scanner and training in keeping the head and body still. MRI and MRS were performed with a 3T Siemens Prisma-fit MRI unit using a 32-channel phased-array head receiver coil with a body transmitter coil. Participants were monitored by video camera located inside the MRI unit and encouraged to hold still regularly.

Magnetization-prepared rapid gradient-echo T1-weighted (T1w) three-dimensional volume imaging was performed with a custom pulse-sequence prepared at the University of Minnesota for the Human Connectome Project using a sagittal multi-echo method that included real-time motion correction with repetition-time (TR)/echo-times (TEs)/inversion-time (TI) = 2500/1.81, 3.6, 5.39, 7.18/1000 ms. The flip angle was 8° and voxel dimensions were 0.8 x 0.8 x 0.8 mm^3^. This acquisition produced a T1w 3-dimensional volume image that was used for positioning of subsequent MRI and ^1^H MRS acquisitions, tissue-segmentation, and radiologic review.

GABA+ J-difference-edited MRS without macromolecule suppression was performed using the manufacturer’s unmodified investigational 859G Works-In-Progress pulse-sequence. This pulse-sequence collects two spectra in an interleaved fashion from a defined volume-of-interest (VOI). One spectrum, the INVERT spectrum, is obtained using a frequency-selective radiofrequency pulse that perturbs the GABA nuclear quantum mechanical system. The second spectrum, the control spectrum (CNTL), is collected without perturbing the GABA system. The difference (DIFF) spectrum is then obtained by vector subtraction of the two spectra. Figure 1 illustrates typical spectra. The DIFF spectrum shows signals in the vicinity of 3.0 ppm that are produced by GABA and the MMBL. The CNTL spectrum is similar to a conventional point-resolved spectroscopy (PRESS) spectrum and features signals produced by many brain metabolites, including Glu. The VOI (single MRS voxel; Figure 2) was positioned to overlap with the anterior midline cerebral cortex (aMCC), as in our prior work (O’Neill et al., 2016,2023). The voxel size was 4.0 cm (anterior-posterior) x 3.0 cm (left-right) x 1.0 cm (superior-inferior) = 12.0 cc. The pulse-sequence timing parameters were TR/TE=2000/68 ms. Two consecutive acquisitions (each of which produces INVERT, CNTL and DIFF spectra) were performed with identical voxel size and positioning. A water-suppressed acquisition used 128 averages to assay the metabolite signals (i.e., GABA and Glu) in a total runtime of 256 sec. A not-water-suppressed (NWS) acquisition was also performed using 8 averages (total runtime 16 s) to assay the water signal. NWS spectra were used to reference metabolite levels to water. Acquired spectra were visually reviewed for artifacts (subtraction errors, phase errors, large lipid signal, motion) to identify 34 high-quality acquisitions from unique subjects.

**Figure 1.**
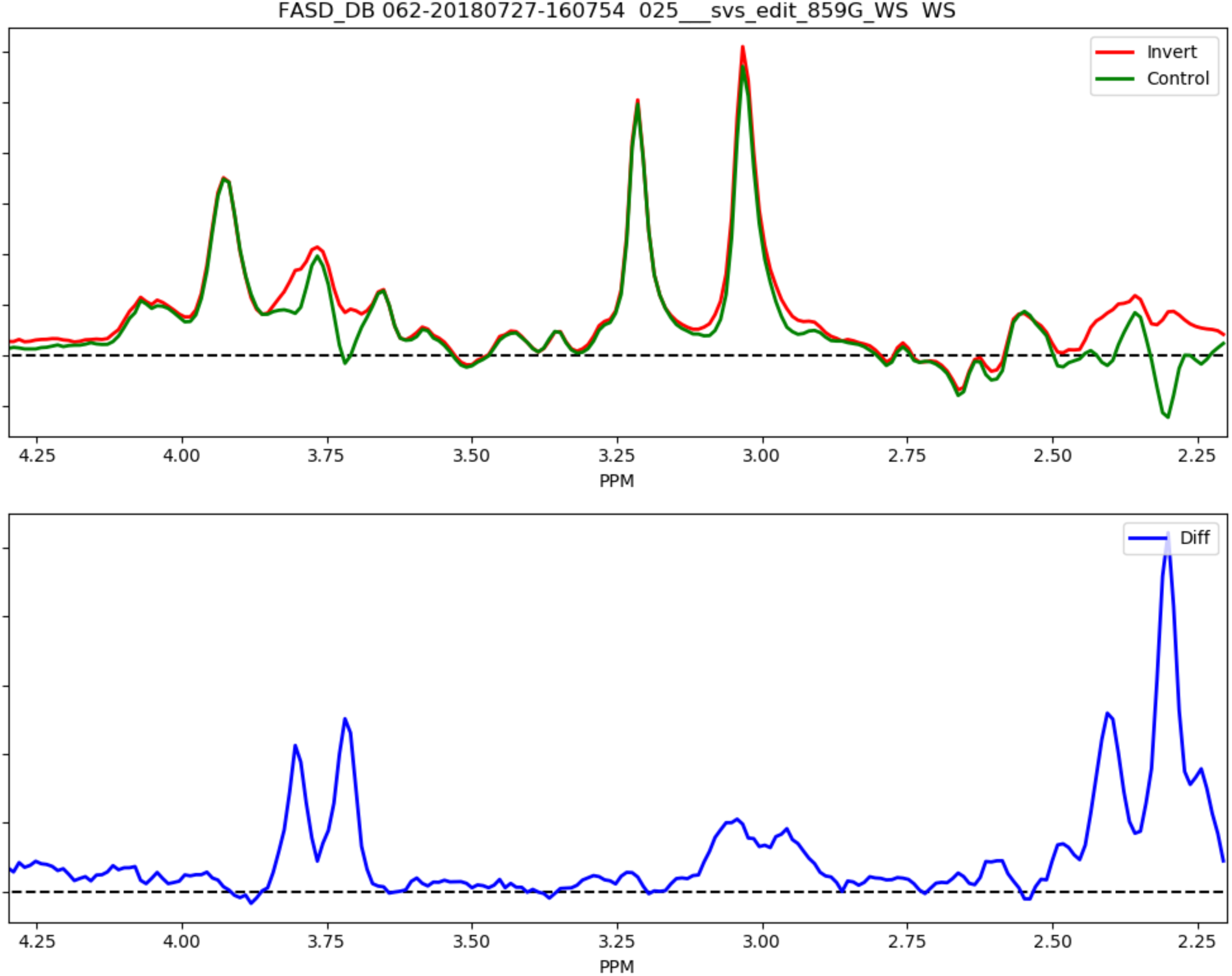
Representative spectra (intensity vs. chemical shift plots) acquired at 3 T with J-difference-editing single-voxel magnetic resonance spectroscopy (MRS; TR/TE=2000/68 ms) of the anterior middle cingulate cortex (aMCC) in children. (*Upper*) the control (CNTL) spectrum (*green*) is like a conventional PRESS spectrum. In the Invert spectrum, the resonance around 3.0 ppm represents GABA+. In offline processing, GABA+ is segregated into GABA and its macromolecular baseline (MMBL).

**Figure 2.**
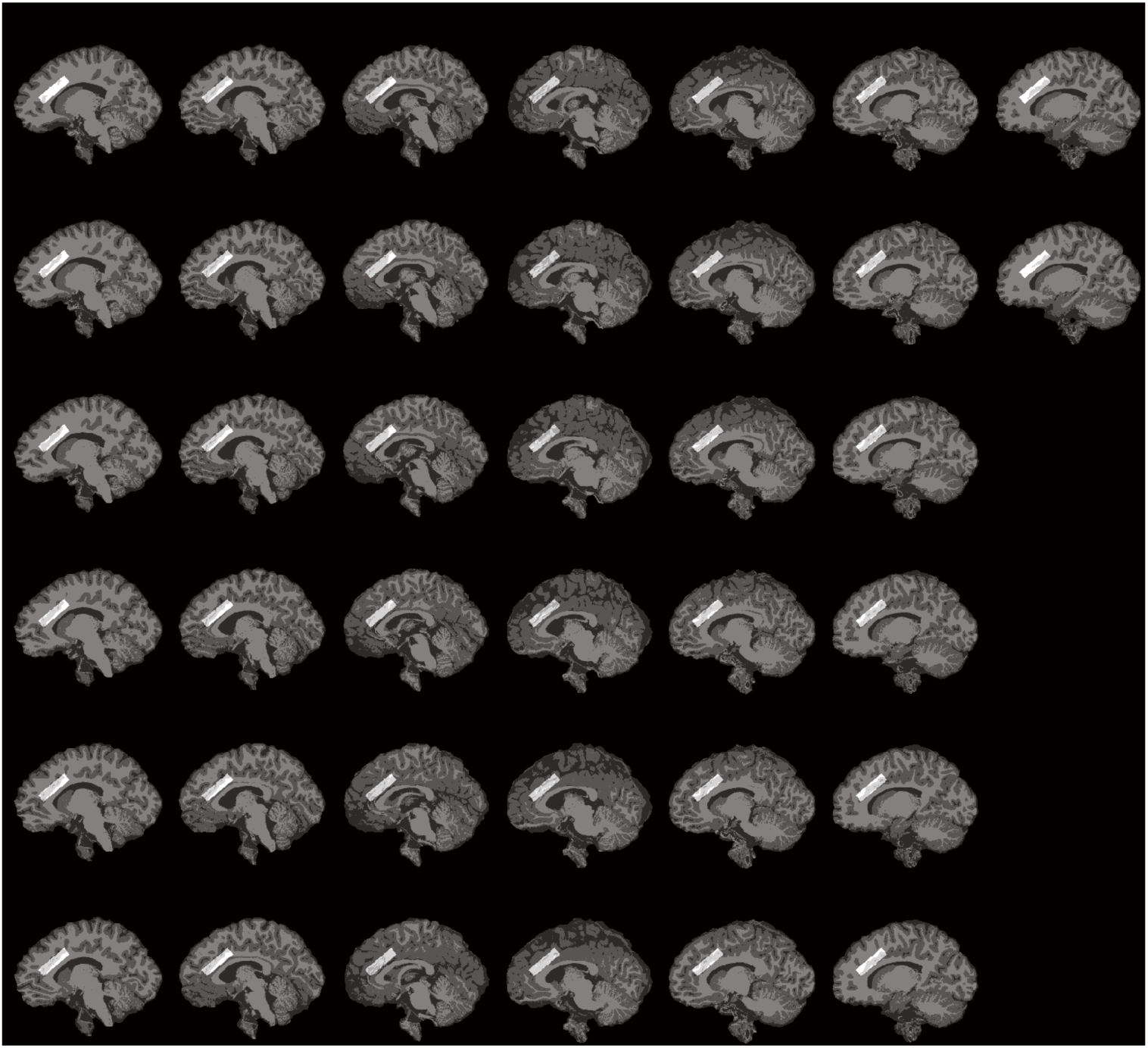
Serial parasagittal section of T1w MRI of a representative study participant’s brain showing location of the J-difference-editing MRS single-voxel (*white*) in the anterior middle cingulate cortex (aMCC). The 4 x 3 x 1 cc voxel was positioned as in our prior work (O’Neill et al., 2016,2023). It was oriented parallel to and sat atop the dorsum of the rostral corpus callosum. The superior (axial-obliquely oriented) face bordered on or extended into neighboring mesial superior frontal cortex; the lateral faces bordered on or extended into local white matter; the anterior face bordered on or extended into the neighboring pregenual anterior cingulate cortex (pACC); the posterior face bordered on or extended into the neighboring posterior middle cingulate cortex (pMCC). Voxel dimensions and orientation were adjusted to maximize local gray-matter content.

### MRS processing

T1w volumes were segmented into gray matter (GM), white matter (WM), and cerebrospinal fluid (CSF) subvolumes using FSL v5.0.10 (Analysis Group, FMRIB, Oxford, UK). MRS voxels were overlaid onto the subvolumes using custom-written software to determine the fraction of GM, WM, and CSF within each voxel.

CNTRL and DIFF spectra were subjected to non-linear least squares fitting to obtain relative metabolite signal levels using a version of SVFit2016 that was modified to read J-difference-edited-MRS. SVFit software, written by JRA (Alger et al., 2016), has been used in multiple prior studies (Alger et al., 2021; Donahue et al., 2022; Grodin et al., 2022,2023; O’Neill et al., 2017,2022,2023). SVFit performs non-linear least squares spectral fitting to determine relative metabolite levels. It uses the Levenberg–Marquardt implementation of the Gauss-Newton method to fit spectra in the frequency domain. Fitting of CNTL spectra included models of spectra for the following metabolites: *N*-acetylaspartate, *N*-acetylaspartylglutamate, Glu, glutamine, GABA, creatine+phosphocreatine (Cr), choline compounds, and *myo*-inositol. Model spectra of residual water, lipids, and macromolecules were also included. Model spectra were simulated in Versatile Simulation, Pulses, and Analysis (VESPA) software (Soher et al., 1998,2023; Young et al., 1998). Similarly, the estimated Glu and glutamine levels were summed to form “Glx”. Fitting of DIFF spectra included models of GABA, MMBL, Glx signals appearing at 3.73 ppm and 3.82 ppm, Cho and Cr, which were included to accommodate the possibility of subtraction errors in the region of the GABA signal. (Note that the signal in the CNTL spectrum and not the DIFF spectrum was used to quantify Glx.) The GABA difference signal model was developed using VESPA to simulate CNTL and INVERT GABA signals appearing in the vicinity of 3.0 ppm and then taking the difference. Glx373, Glx382, Cho and Cre signals were simulated using VESPA. The MMBL signal model that overlaps with GABA was extracted from Mikkelsen et al. (2017). The GABA and MMBL model spectra used are presented in Figure 3. Following fitting, GABA and MMBL signals were summed to obtain GABA+. All metabolite levels were water-referenced to the water level obtained by fitting the NWS CNTL water signal. All water-referenced metabolites were then corrected for the voxel CSF-content by dividing the uncorrected levels by 1 – volume fraction CSF. Spectra were rejected with signal-to-noise ratio (SNR) < 5 or linewidth > 0.03 ppm; voxels were rejected with GM < 40% or CSF > 15%. (See Supplemental Materials for MRSinMRS checklist; Lin et al., 2021.)

**Figure 3.**
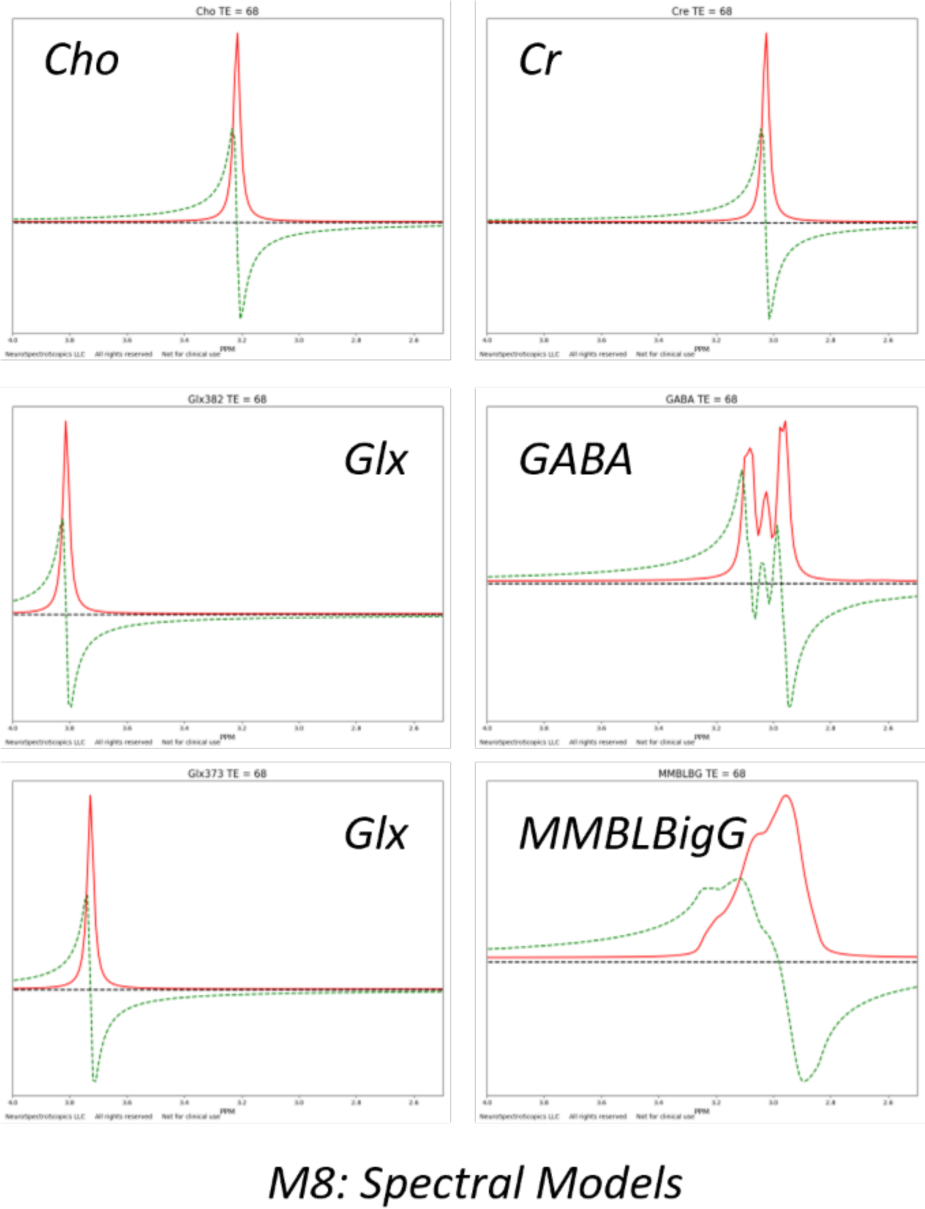
Model spectra for DIFF GABA and MMBL.

### Statistical Analysis

Between-group differences in age were assessed using the nonparametric Kruskal-Wallis (KW) Test. Relationships between each target metabolite level and age and clinical diagnosis were evaluated using general linear ordinary least squares modelling (GLM) provided by statsmodels (www.statsmodels.org). Analysis began by including all three groups (ADHD+PAE, ADHD-PAE, TD) and testing for significance (*p* < 0.05) using the following omnibus model:

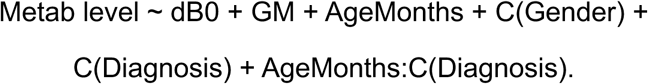

This model posits that the metabolite level depends on the following continuous variables: magnetic field inhomogeneity (dB0) within the VOI, GM voxel content (GM) within the VOI, and participant age (AgeMonths) together with the following categorical variables: Gender (Male or Female), Diagnosis (ADHD+PAE, ADHD-PAE, TD). dB0 is the full-width-at-half maximum fitted CNTL 3.03 ppm Cr signal. It is included in the statistical model to accommodate the potential impact of spectral quality on measured metabolite levels. GM is included to account for likely differences in metabolite levels between GM and WM. AgeMonths is included to detect possible age-related changes in metabolite levels and to accommodate imbalances in age between the three groups. Gender is included to accommodate the imbalance in gender between the three groups. The AgeMonths:C(Diagnosis) is an interaction term that detects possible between-group differences in the effects of participant age on metabolite levels.

Whenever the omnibus model revealed a significant main effect of age, main effect of diagnosis or age-by-diagnosis interaction, *post-hoc* analyses were performed to identify significant effects between each pair of groups (ADHD+PAE versus ADHD-PAE, ADHD+PAE versus TD, ADHD-PAE versus TD) as follows:

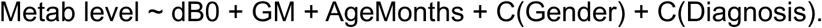

For each main effect or interaction, each omnibus or *post-hoc* GLM yields a fitting coefficient (“coef”) as the corresponding effects size, as well as values for the number of observations (“nobs”), number of parameters (“nparams”), degrees-of-freedom (“dof”), and a *p*-value. The above omnibus approach additionally affords a (modest) protection against multiple comparisons.

## RESULTS

Group-mean values for the target metabolite levels and ratios (Glu, Glx, GABA, GABA/Glu, GABA/Glx, GABA+, MMBL) are listed for the three groups in Table 2, along with the corresponding mean tissue-compositions of the MRS voxel. There were no significant between-group differences in tissue composition (all *p* > 0.05). Effects of age and diagnosis on the target metabolite endpoints are reported below.

**Table 2.** Group-mean metabolite levels in midline anterior middle cingulate cortex (aMCC).

|  | ADHD+PAE | ADHD-PAE | TD |
| --- | --- | --- | --- |
| **GM**, volume% | 54.0±4.7 | 55.3±5.8 | 56.8±4.0 |
| **WM**, volume% | 37.6±8.5 | 36.5±7.1 | 34.3±5.3 |
| **CSF**, volume% | 8.5±2.6 | 8.2±2.4 | 8.9±2.7 |
| **Glu**, IU | 20.5±2.7 | 20.4±3.0 | 21.7±2.5 |
| **Glx**, IU | 22.9±2.9 | 22.7±3.1 | 24.2±2.5 |
| **GABA**, IU | 2.0±0.9 | 2.1±1.1 | 1.9±0.9 |
| **GABA/Glu**, dimensionless | 0.10±0.05 | 0.11±0.06 | 0.09±0.04 |
| **GABA/Glx**, dimensionless | 0.09±0.04 | 0.09±0.05 | 0.08±0.04 |
| **GABA+**, IU | 11.2±2.0 | 11.0±2.9 | 11.8±2.8 |
| **MMBL**, IU | 9.2±2.6 | 8.9±3.8 | 9.9±3.1 |

ADHD+PAE=attention deficit hyperactivity disorder likely due to prenatal alcohol exposure; ADHD-PAE=attention deficit hyperactivity disorder without prenatal alcohol exposure, likely due to genetics; TD=typically developing control. GM=voxel gray-matter content; WM=voxel white-matter content; CSF=voxel cerebrospinal fluid content; Glu=glutamate; Glx=glutamate+glutamine; GABA= $\gamma$ -aminobutyric acid; GABA= $\gamma$ -aminobutyric acid+ macromolecular baseline; MMBL=macromolecular baseline; IU=Institutional Units; \* $p < 0.05$ , \*\*\* $p < 0.001$ vs. ADHD-PAE; † $p < 0.05$ , †† $p < 0.001$ vs. TD. Glu and Glx are fit from the CNTL spectra; GABA, GABA+ and MMBL from the DIFF spectra.

### Effects of Age and Diagnosis on Glu and Glx

Omnibus GLM analyses did not reveal any significant main effects of diagnosis or age-by-diagnosis interactions for Glu or Glx (all *p* > 0.05).

### Effects of Age and Diagnosis on GABA, GABA/Glu, and GABA/Glx

Omnibus analysis for GABA found a significant main effect of diagnosis (coef = 0.033, nobs = 76, nparams = 8, dof = 67, *p* = 0.025). Thereby, GABA was higher in ADHD+PAE and ADHD-PAE than in TD. A significant age-by-diagnosis interaction (coef = −4.105, nobs = 76, nparams = 8, dof = 67, *p* = 0.024) also resulted. In *post-hoc* analyses, GABA increased significantly with increasing age for TD (coef = 0.019, nobs = 28, nparams = 4, dof = 23, *p* = 0.022), but not for ADHD+PAE or ADHD-PAE (both *p* > 0.05).

Results for GABA/Glu and GABA/Glx closely matched those for GABA. For GABA/Glu, omnibus analysis found a significant main effect of diagnosis (coef = −0.231, nobs = 76, nparams = 8, dof = 67, *p* = 0.018), whereby GABA/Glu was higher for ADHD+PAE and ADHD-PAE than for TD. Additionally, the age-by-diagnosis interaction for GABA/Glu was significant (coef = 0.002, nobs = 76, nparams = 8, dof = 67, *p* = 0.018). In *post-hoc* analysis, GABA/Glu increased with increasing age for TD (coef = 0.001, nobs = 28, nparams = 4, dof = 23, *p* = 0.012) but not for ADHD+PAE or ADHD-PAE (both *p* > 0.05). For GABA/Glx, omnibus analysis found a significant main effect of diagnosis (coef = 0.028, nobs = 76, nparams = 8, dof = 67, *p* = 0.027), whereby GABA/Glx was higher for ADHD+PAE and ADHD-PAE than for TD. The age-by-diagnosis interaction for GABA/Glx was significant (coef = 0.027, nobs = 76, nparams = 8, dof = 67, *p* = 0.028). *Post-hoc*, GABA/Glx increased with increasing age for TD (coef = 0.001, nobs = 28, nparams = 4, dof = 23, *p* = 0.025) but not for the two ADHD groups (both *p* > 0.05).

### Effects of Age and Diagnosis on GABA+ and MMBL

Effects observed for GABA+ (i.e., GABA including MMBL) did not recapitulate those seen for GABA there being no significant main effect of diagnosis or age-by-diagnosis interaction (both *p* > 0.05). Omnibus testing did reveal a significant main effect of age (coef = 0.059, nobs = 76, nparams = 8, dof = 67, *p* = 0.049). *Post-hoc*, GABA+ increased with increasing age for ADHD+PAE (coef = 0.056, nobs = 27, nparams = 4, dof = 22, *p* = 0.024) but not for ADHD-PAE or TD (both *p* > 0.05).

Finally, the macromolecular baseline (MMBL) exhibited a significant main effect of age (*p* = 0.034) for the ADHD+PAE group (coef = 0.071, nobs = 27, nparams = 4, dof = 22, *p* = 0.034) but not for the ADHD-PAE and TD groups (both *p* > 0.05).

## DISCUSSION

To our knowledge, this is the first comparison of MRS GABA in children with ADHD associated with prenatal alcohol exposure (ADHD+PAE) versus non-exposed children with probable familial ADHD (ADHD-PAE). There were three major findings: 1) In anterior middle cingulate cortex (aMCC), levels of GABA were near equal in ADHD+PAE and ADHD-PAE and were both higher than in TD; 2) aMCC GABA increased with age in TD, but not in ADHD+PAE or ADHD-PAE; and 3) Results for GABA+, i.e., GABA not isolated from its macromolecular spectral background, in aMCC did not mirror results for GABA itself. Taken together, these findings imply that abnormal level and age-trajectory of aMCC GABA are features common to ADHD of at least two etiologies: familial inheritance and *in utero* ethanol exposure. Additionally, caution is advised in interpreting GABA+ measures from J-difference-edited studies.

The first major finding was that aMCC GABA did not differ significantly between ADHD+PAE and ADHD-PAE. In past work our group identified multiple neuroimaging markers that distinguished ADHD+PAE from ADHD-PAE or even enabled classification of individual patients. These measures included: Cho and fractional anisotropy (FA) in anterior corona radiata (O’Neill et al., 2019); Glu, FA and diffusivity indices, especially in frontal white matter (Alger et al., 2021; Kilpatrick et al. 2022; O’Neill et al., 2022); local gyrification index (Kilpatrick et al., 2021) and intracortical myelin (Kilpatrick et al., 2022) in widespread cortex; and resting-state fMRI functional connectivity within the frontoparietal network (FPN; Gupta et al., submitted). Based on present data, GABA in the aMCC is *not* a measure that distinguishes ADHD+PAE, rather it appears to indicate a commonality between the two etiologies. Thus, the aMCC may be one structure in a “final common pathway” in the brain that brings about ADHD as a syndrome or, alternatively, a downstream structure that is altered once one has ADHD.

After taking account of age, gender, tissue-composition, and spectral quality, aMCC GABA was above normal for both conditions. The same was seen for GABA/Glu and for GABA/Glx. No significant effects of diagnosis were observed for Glu or Glx. This suggests that ADHD is influenced by GABAergic, rather than glutamatergic, abnormalities (at least in the aMCC). Thus, present findings perhaps represent excess GABA-related inhibition in the aMCC which is not counterbalanced by Glu-related excitation. This may be consistent with a proposal we made previously (O’Neill et al., 2013) that ADHD is characterized by an “overdamping” (over-inhibition) of glutamatergic activity in cortex. Since most GABAergic projections in the cerebral cortex are local, the excess GABA producing this putative enhanced inhibition is possibly synthesized within the aMCC. This inhibition may hamper the excitatory output of the aMCC to other brain regions and thus impair the execution of aMCC cognitive functions, including allocation of attention.

Relevant literature findings are somewhat sparse. The closest comparisons to the present study are Ende et al. (2018) and Hai et al. (2020). Hai et al. (2020) found no effects of pediatric ADHD on GABA, Glu, or Glx in aMCC, but their brief publication (a letter) does not make clear how the MMBL was handled in their study. Using online suppression of MMBL, as opposed to the offline removal of MMBL used here, Ende et al. (2018) observed not above-, but below-normal GABA in ADHD. That, however, was in an adult sample.

Investigations in other brain regions imply that GABA may differ in adult versus pediatric ADHD (Bollmann et al. 2015) and that stimulant treatment initiated in childhood versus in adulthood may reduce GABA+ in adult patients (Solleveld et al., 2017). At 7 T in pediatric ADHD, Puts et al. (2020) measured GABA/Cr without spectral-editing in a voxel containing some aMCC, but mainly neighboring pregenual anterior cingulate cortex (pACC). No effects of ADHD were found. The groundbreaking study of Edden et al. (2012) measured lower GABA in children with ADHD than in TD children in left sensorimotor cortex. Metabolite levels were adjusted for MMBL with an offline correction factor. The same group in the same brain region (Harris et al. 2021), found no difference in GABA+ between ADHD and TD children. They called this a “failure to replicate”, but that is inexact, given that the later study did not correct for MMBL. Given the paucity and heterogeneity of methods in the literature to date, it is difficult to judge the extent of agreement of present findings with prior results. More exact replicative studies are called for.

The second major finding was that aMCC GABA increased with age in TD, but not in ADHD+PAE or ADHD-PAE. This was also seen for GABA/Glu and GABA/Glx in the absence of age effects on Glu or Glx. In other words, there is reasonable (albeit cross-sectional) evidence that the age trajectory of aMCC GABA in ADHD of either etiology differs from normal. As further mean increase of GABA with age was not evident in the ADHD groups, excess GABA in ADHD may be self-limiting. Longitudinal studies of GABA in ADHD could better evaluate the possible presence of these abnormal age-trajectories.

The third major finding was that the above-discussed effects of diagnosis and of age on GABA were not evident when GABA+ was analyzed in place of GABA. Instead, an increase in GABA+ with age was seen not for the TD but for the ADHD+PAE group only. It is plausible that this GABA+ age-increase was driven by the increase of MMBL observed with age, which was similarly limited to ADHD+PAE. This is especially likely as the magnitude of the MMBL signal was actually larger than that of GABA. In any case, present results indicate that failure to account for the confounding impact of MMBL could be problematic when assessing effects of age and diagnosis on GABA. Our fitting method could be applied to MR spectra acquired in other disorders to determine how closely GABA and GABA+ track together in those conditions.

A minor finding was the absence of effects of age or diagnosis on Glu or Glx. These results should be accepted with caution since present data were acquired at TE=68 ms, a value that may be suboptimal for detection of Glu.

This study has several limitations. Sample size was modest (20-30 subjects per group); replication in larger samples is called for. As mentioned above, the ADHD+PAE sample was younger, more heavily medicated, and had lower FSIQ than the other two samples. Several ADHD+PAE participants, and none in the other groups, also had other teratogenic exposures beyond ethanol. These characteristics are unfortunately typical of PAE populations. Age-of-onset of ADHD symptoms is younger in alcohol-exposed than in unexposed children (O’Malley & Nanson, 2002; Peadon et al., 2010). Since they are often refractory to conventional stimulant medication, children with ADHD+PAE are frequently treated with heavier doses and polypharmacy. Low IQ is a common symptom of PAE. Finally, it is common for women who drink while pregnant to consume other substances-of-abuse as well. These complications render it challenging to recruit well-matched samples along these dimensions. As mitigating factors, age was included in all statistical models of metabolite effects, children were assessed while off stimulant medications, all participants had FSIQ in the normal range (>70), and ethanol was the dominant (in many cases only) prenatal exposure for all ADHD+PAE participants. Limitations of the J-edited-MRS acquisition include its large voxel size which can foster partial-voluming. This was accounted for by correcting for voxel CSF-content and by including voxel GM in the model. Notwithstanding these limitations, present findings point to a possible abnormal excess of GABA in two etiologies of ADHD in a cortical region long implicated in the disorder.

## Data Availability

All data produced in the present study are available upon reasonable request to the authors

## SUPPLEMENTAL MATERIALS

**Table S1.** MRSinMRS checklist.

| Site (Name or Number) |  |
| --- | --- |
| 1. Hardware |  |
| a. Field strength [T] | 3 T |
| b. Manufacturer | Siemens |
| c. Model (software version if available) | Prisma VE11C |
| d. RF coils: nuclei (transmit/receive), number of channels, type, body part | proton, 32 channels, phased-array, head |
| e. Additional hardware | none |
| 2. Acquisition |  |
| a. Pulse sequence | Siemens 839G WIP<br>J-difference-edited single-voxel MRS |
| b. Volume of Interest (VOI) locations | midline (left+right)<br>anterior middle<br>cingulate cortex |
| c. Nominal VOI size [cm<sup>3</sup>, mm<sup>3</sup>] | 4.0 cm (anterior-posterior) x 3.0 cm (left-right) x 1.0 cm (superior-inferior) = 12.0 cc |
| d. Repetition Time (TR), Echo Time (TE) [ms, s] | 2000/68 ms |
| e. Total number of Excitations or acquisitions per spectrum | 128 Invert/128 Control |
| In time series for kinetic studies |  |
| i. Number of Averaged spectra (NA) per time-point |  |
| ii. Averaging method (e.g. block-wise or moving average) |  |
| iii. Total number of spectra (acquired / in time-series) |  |
| f. Additional sequence parameters<br>(spectral width in Hz, number of spectral points, frequency offsets)<br>If STEAM: Mixing Time (TM)<br>If MRSI: 2D or 3D, FOV in all directions, matrix size, acceleration factors, sampling method | 1200 Hz, 1024 spectral points, delta -2.8 ppm<br>edit pulse frequency 1.90 ppm<br>edit center frequency 4.70 ppm |
| g. Water Suppression Method | CHESS |
| h. Shimming Method, reference peak, and thresholds for “acceptance of shim” chosen | Siemens Brain shim |
| i. Triggering or motion correction method<br>(respiratory, peripheral, cardiac triggering, incl. device used and delays) | none |
| **3. Data analysis methods and outputs** |  |
| a. Analysis software | SVFit2016 |
| b. Processing steps deviating from quoted reference or product | none |
| c. Output measure<br>(e.g. absolute concentration, institutional units, ratio)Processing steps deviating from quoted reference or product | Institutional Units, ratios |
| d. Quantification references and assumptions, fitting model assumptions | water reference |
| **4. Data Quality** |  |
| a. Reported variables<br>(SNR, Linewidth (with reference peaks)) | SNR, linewidth of Cr 3.03 ppm peak |
| b. Data exclusion criteria | SNR<5,<br>Linewidth>0.03<br>voxel GM < 40% |
|  | voxel CSF > 15% |
| c. Quality measures of postprocessing Model fitting (e.g. CRLB, goodness of fit, SD of residual) | CRLB |
| d. Sample Spectrum | included |

